# Artificial Intelligence Agent: AI-HOPE-TP53 Enables Pathway-Centric Analysis of TP53-Driven Molecular Alterations in Colorectal Cancer Precision Oncology

**DOI:** 10.1101/2025.05.23.25328251

**Authors:** Ei-Wen Yang, Brigette Waldrup, Enrique Velazquez-Villarreal

## Abstract

**Introduction:** Early-onset colorectal cancer (EOCRC) is rising rapidly, especially among populations at risk who experience disproportionate incidence and mortality. The TP53 pathway, frequently altered in CRC, regulates key processes such as DNA repair and apoptosis. Despite its clinical relevance, TP53 dysregulation remains understudied in EOCRC, particularly in populations at risk. Current tools lack support for pathway-specific, population-stratified, and treatment-aware analysis. To address this gap, we developed AI-HOPE-TP53—the first conversational AI agent designed to explore TP53 alterations in CRC using harmonized clinical-genomic data and natural language queries.

**Methods:** AI-HOPE-TP53 leverages large language models (LLMs) to translate natural language prompts into executable bioinformatics workflows operating on datasets from cBioPortal. It supports dynamic cohort stratification by TP53-related genes, age, MSI status, treatment, tumor stage, and ethnicity. Automated analyses include mutation frequency, co-mutation profiling, survival curves, and odds ratios.

**Results:** AI-HOPE-TP53 successfully replicated known molecular characterization, identifying higher TP53 alteration rates in EOCRC patients. It revealed trends toward improved survival in patients with TP53 mutations and identified significant enrichment of H/L individuals among early-onset, FOLFOX-treated cases (OR = 2.002, p < 0.0001). Additional analyses uncovered subsite-specific outcomes in ATM-mutant CRC, stage-specific effects in CHEK1-mutated tumors, and disproportionate health burdens in treatment exposure.

**Conclusions:** AI-HOPE-TP53 enables fast, natural language–driven exploration of TP53 pathway alterations in CRC. Its ability to integrate clinical and genomic data across diverse subgroups makes it a powerful tool for precision oncology and Community-engaged health research—supporting both hypothesis validation and discovery in EOCRC and beyond.

## Introduction

Colorectal cancer (CRC) remains the third most commonly diagnosed cancer and the second leading cause of cancer-related deaths worldwide [1–3]. While the overall incidence has stabilized in high-income countries, early-onset CRC (EOCRC)—defined as CRC diagnosed before the age of 50—has emerged as a rapidly increasing subset with rising incidence and mortality rates [4–7]. This upward trend is especially pronounced in Hispanic/Latino (H/L) populations, who now exhibit the highest increase in both EOCRC incidence and mortality compared to non-Hispanic Whites (NHW) [8–11]. These disproportionate health burdens call for deeper exploration of genomic alterations that may uniquely affect EOCRC in underrepresented populations.

Notably, EOCRC often presents at more advanced stages, likely due to the absence of routine screening before age 50 [12]. Moreover, molecular studies have revealed that EOCRC is biologically distinct from late-onset CRC (LOCRC), with higher rates of microsatellite instability (MSI), tumor mutation burden (TMB), and PD-L1 expression, as well as epigenetic changes like LINE-1 hypomethylation [13–15]. Among the most critical molecular drivers of CRC are alterations in the TP53 signaling pathway, which influence tumor behavior, progression, and therapeutic response.

The TP53 pathway governs a broad array of essential cellular processes including DNA repair, apoptosis, autophagy, metabolism, and cell cycle regulation. The tumor suppressor protein p53—often referred to as the “guardian of the genome”—is central to this network, initiating transcriptional programs that ensure genomic integrity following DNA damage through genes such as CDKN1A (p21), GADD45, and BAX [16–18]. TP53 is mutated in approximately 74% of CRC tumors, representing one of the most frequently altered genes in this malignancy [19–22]. Furthermore, missense gain-of-function mutations in TP53 are increasingly linked to therapeutic resistance and poor clinical outcomes [23–26]. Despite its well-established role in cancer, the specific contribution of TP53 pathway alterations to EOCRC—especially among H/L patients— remains under-characterized, limiting efforts to develop population-specific therapeutic strategies [19, 23,27,28].

Current analysis platforms such as cBioPortal [29] and UCSC Xena [30] offer access to large-scale datasets including TCGA and AACR GENIE, yet they lack user-friendly, pathway-focused tools that can simultaneously integrate genomic and clinical parameters with contextual attention to race/ethnicity, age, and treatment exposure. These limitations hinder precision oncology efforts, particularly in real-world and underserved populations. In addition, non-computational researchers face barriers in performing integrative analyses across multi-omic, clinical, and disparity-focused dimensions, leaving gaps in the translation of genomic data into actionable insights. Recent advances in artificial intelligence (AI), specifically through LLMs, offer transformative potential for addressing these gaps by enabling natural language–driven bioinformatics [31–35]. Emerging conversational AI platforms can automate hypothesis generation, survival modeling, and stratified cohort analysis from harmonized datasets—yet few are designed to interrogate pathway-specific events such as TP53 dysregulation or tailored to support disparity-aware research in CRC.

To address this critical need, we developed AI-HOPE-TP53 (Artificial Intelligence agent for High-Optimization and Precision Medicine focused on TP53), a first-in-class conversational AI system purpose-built for pathway-specific, integrative CRC analysis. AI-HOPE-TP53 allows users to pose natural language questions that trigger backend computational workflows, automating survival analysis, odds ratio estimation, and population disaggregation across clinical and genomic variables. In this study, we: (1) developed AI-HOPE-TP53 to facilitate user-friendly exploration of TP53 pathway alterations in CRC; (2) validated its performance through replication of known genotype-phenotype associations; and (3) applied it to uncover novel links between TP53 mutations, MSI status, tumor stage, and ethnicity in EOCRC. These findings demonstrate AI-HOPE-TP53 as a scalable and inclusive platform for advancing precision oncology through pathway-centered, population-aware bioinformatics.

## Methods

### Architecture and operational framework of AI-HOPE-TP53

AI-HOPE-TP53 is a conversational artificial intelligence (AI) platform purpose-built to investigate the clinical and genomic implications of TP53 pathway alterations in colorectal cancer (CRC). The system integrates four functional modules: (1) a LLM interface for natural language comprehension, (2) a query translation engine that converts user prompts into programmatic instructions, (3) a filtering layer for cohort construction based on clinical-genomic constraints, and (4) a statistical processing engine for automated analysis and visualization (Figure 1). Upon receiving a query— such as “Compare survival between TP53-mutant and wild-type tumors in Hispanic/Latino EOCRC patients”—the LLM parses semantic intent, activates a corresponding backend pipeline, and returns outputs including Kaplan-Meier plots, odds ratios, and interpretive summaries.

**Figure 1.**
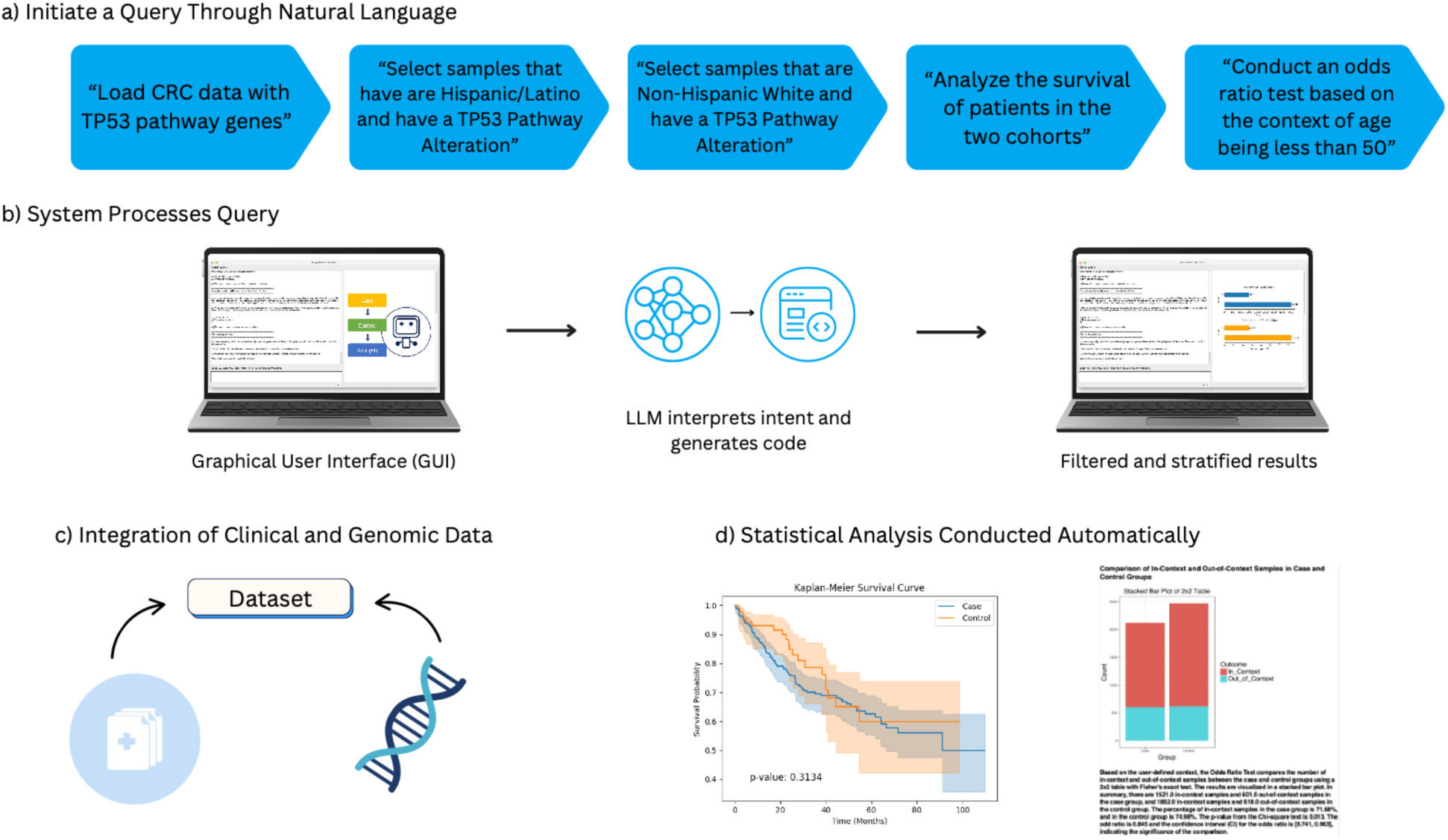
Schematic representation of AI-HOPE-TP53 query and analysis pipeline. This figure outlines the core operational stages of AI-HOPE-TP53, a conversational AI agent designed to analyze TP53-related genomic events in colorectal cancer (CRC) through integrative, automated workflows. a) The process begins with a series of sequential natural language inputs, where users specify analysis parameters such as cohort definitions (e.g., TP53-altered Hispanic/Latino vs. Non-Hispanic White patients), survival outcome comparisons, or subgroup-specific odds ratio testing for early-onset cases. b) These inputs are parsed by a graphical user interface (GUI) that channels them through a back-end engine, where a LLM translates user intent into code-based instructions and dynamically builds filtered clinical-genomic subsets. c) AI-HOPE-TP53 connects to curated datasets—including cBioPortal—and extracts relevant molecular and phenotypic information centered on TP53 and its associated regulatory genes (e.g., CDKN1A, GADD45A, MDM2, BAX, ATM). d) Once executed, the platform returns fully stratified results and automated statistical outputs, including Kaplan-Meier survival curves, contingency tables, and effect size visualizations.

### Data integration and curation

To support its pathway-centric analyses, AI-HOPE-TP53 integrated: (1) harmonized multi-omic datasets from the public repository cBioPortal [29]; (2) genomic profiling data focused on TP53 and associated regulators such as CDKN1A, GADD45A, MDM2, BAX, and ATM [2]; (3) comprehensive clinical annotations, including patient age, race/ethnicity, tumor location, disease stage, MSI status, treatment history, and survival outcomes; and (4) standardized preprocessing pipelines that ensured analytical consistency through ontology normalization (e.g., OncoTree), sample ID harmonization, and conversion into tabular, analysis-ready matrices.

### Query interpretation and dynamic cohort construction

The core AI engine of AI-HOPE-TP53 employed a fine-tuned LLaMA 3–based LLM trained for biomedical domain specificity. It enabled flexible interpretation of natural language queries and supported dynamic cohort generation across intersectional variables. For example, prompts like “Analyze TP53 mutation rates in high TMB H/L patients under 50” were translated into multi-layered filters encompassing age, mutation status, race/ethnicity, and tumor mutation burden. The platform also allowed cohort stratification by TP53 mutation type (e.g., missense, nonsense, truncating), microsatellite instability (MSI) status, tumor site (primary vs. metastatic), age group (<50 vs. ≥50 years), and racial/ethnic categories (e.g., H/L vs. NHW).

### Statistical framework and analytical capabilities

Statistical analyses within AI-HOPE-TP53 were implemented in Python, utilizing libraries such as Lifelines, SciPy, and StatsModels. For categorical comparisons (e.g., mutation frequency by ethnicity), the system applied chi-square or Fisher’s exact tests. Odds ratios and 95% confidence intervals were automatically calculated to quantify effect sizes. For survival analysis, the platform used Kaplan-Meier estimators with log-rank tests to assess differences between groups. Cox proportional hazards models were employed to adjust for confounding in multivariable settings. Additional modules support the interrogation of TP53 co-mutations (e.g., *MDM2* or *BAX*) and analysis of subpopulation-level – disproportionate health burdens [2].

### Model validation and reproducibility strategy

To ensure analytical validity and reproducibility, AI-HOPE-TP53 incorporates a retrieval-augmented generation (RAG) system that cross-references structured biomedical content. Query interpretation is schema-guided, reducing output variability and minimizing hallucination risk. The system was validated by replicating previously reported genotype-phenotype associations involving TP53 mutations, such as their link to poor survival outcomes and elevated mutation frequencies in H/L EOCRC patients [2].

### Benchmarking and user experience evaluation

Test cases included ethnicity-stratified mutation analysis, MSI-specific survival curves in TP53-mutant CRC, and co-occurrence mapping for TP53 and CDKN1A. AI-HOPE-TP53 demonstrated enhanced usability, faster query resolution, and greater flexibility in defining intersectional clinical-genomic cohorts.

### Visualization and export functions

All outputs from AI-HOPE-TP53 were paired with high-resolution visualizations generated using Plotly and Matplotlib, including Kaplan-Meier survival curves, bar charts, forest plots for odds ratios, and co-mutation heatmaps. Each analysis was accompanied by narrative summaries contextualized within current CRC literature. Results were exportable in multiple formats (CSV, PNG, PDF) to support seamless integration into manuscripts, clinical reports, and presentations.

## Results

### Application of AI-HOPE-TP53 to pathway-centric and disparity-aware analyses

To evaluate the performance, scalability, and translational relevance of AI-HOPE-TP53, we implemented a series of natural language–driven analyses across multiple clinical and molecular dimensions of colorectal cancer (CRC). These analyses were designed to validate the system’s ability to replicate known associations between TP53 pathway alterations and clinical outcomes, while also assessing its potential to uncover novel insights at the intersection of genomics, treatment exposure, and disproportionate health burdens. Using harmonized datasets from cBioPortal, AI-HOPE-TP53 enabled fully automated case-control stratification, survival modeling, and statistical testing based on conversational queries. Here we present the results of these analyses, starting with validation tasks that benchmarked AI-HOPE-TP53 against known genotype-phenotype patterns, followed by exploratory investigations highlighting its utility in hypothesis generation.

### Validation analysis: TP53 pathway alterations in EOCRC by ethnicity

We first assessed AI-HOPE-TP53’s ability to detect ethnicity-based differences in TP53 pathway alterations among early-onset colorectal cancer (EOCRC) patients. Among those with colon adenocarcinoma and age <50, H/L patients showed a higher prevalence of TP53 pathway alterations (91.46%) compared to NHW patients (83.39%), yielding an odds ratio of 2.13 (95% CI: 0.956–4.767; p = 0.084) (Figure 2). While not statistically significant, the trend was suggestive of a potential enrichment in the H/L group.

**Figure 2.**
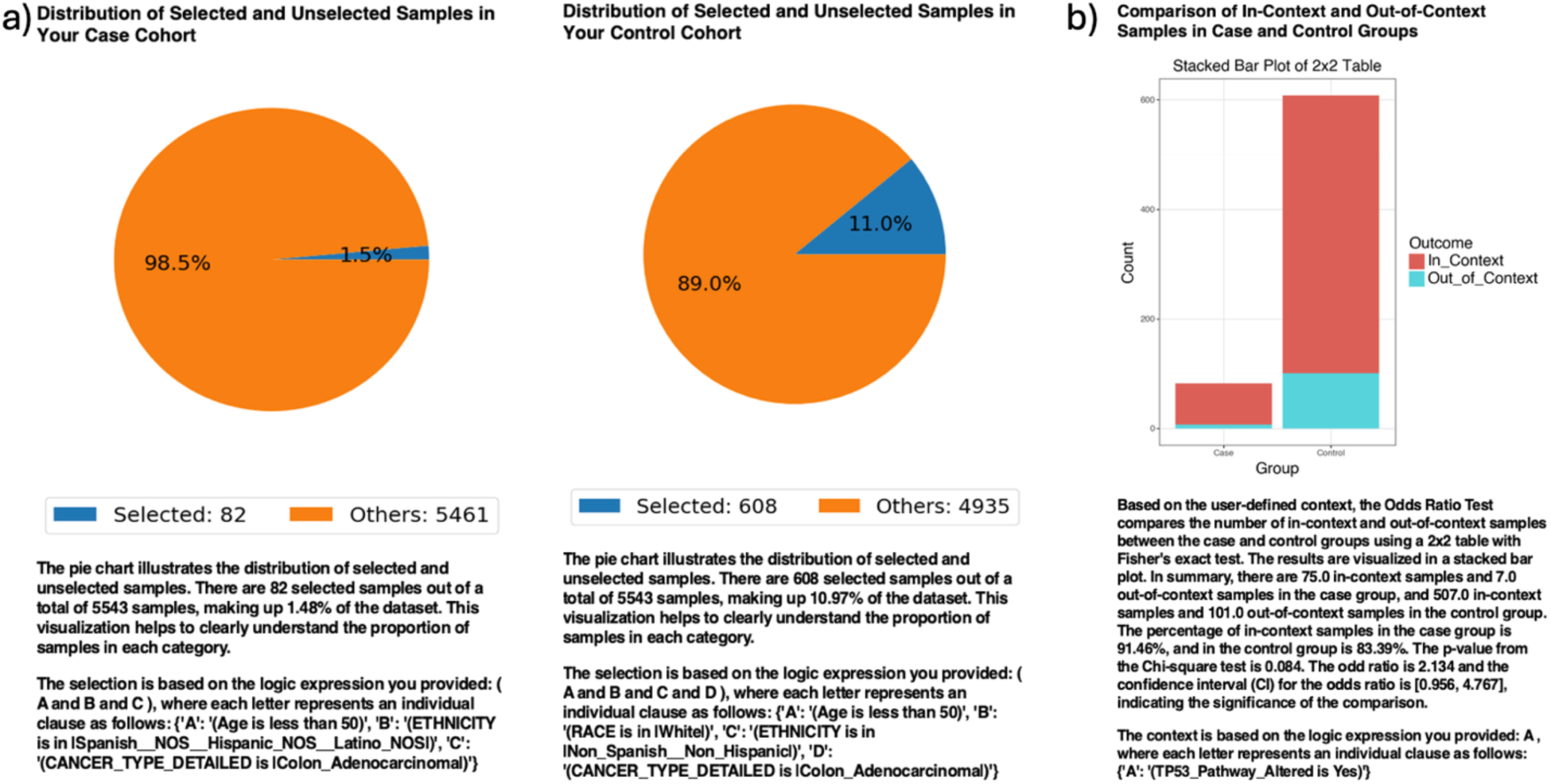
AI-HOPE-TP53 analysis of TP53 pathway alterations in early-onset colon adenocarcinoma among Hispanic/Latino (H/L) vs. Non-Hispanic White (NHW) patients. a) Pie charts illustrate the distribution of selected samples following natural language–driven filtering of the dataset. The case cohort includes 82 early-onset colorectal cancer (EOCRC) patients under age 50 with H/L ethnicity and a diagnosis of colon adenocarcinoma, comprising 1.5% of the dataset. The control cohort includes 608 EOCRC patients under age 50 who are NHW, and diagnosed with colon adenocarcinoma, representing 11% of the dataset. b) A 2×2 odds ratio analysis evaluates the frequency of TP53 pathway alterations between the two groups. The stacked bar plot shows the proportion of samples with and without TP53 pathway alterations. Alterations were present in 91.46% of H/L cases and 83.39% of NHW controls. The calculated odds ratio was 2.13 (95% CI: 0.956–4.767), with a p-value of 0.084, indicating a non-significant but suggestive trend toward greater TP53 alteration prevalence in the H/L population.

When broadening the inclusion criteria to all EOCRC patients regardless of tumor subsite, similar findings emerged. In this larger cohort, TP53 pathway alterations were found in 90.2% of H/L patients versus 85.05% in NHW patients, corresponding to an odds ratio of 1.62 (95% CI: 0.926–2.825; p = 0.114) (Figure S1). These results validate the system’s ability to replicate known disproportionate health burdens and emphasize its capability to handle ethnic stratification in pathway-specific contexts.

### Exploratory analysis: Ethnicity-specific survival outcomes in TP53-Mutant CRC

To examine whether ethnicity modulates survival in TP53-mutant CRC, we analyzed primary tumor cases stratified by H/L and NHW background. The Kaplan-Meier survival analysis revealed a non-significant trend toward improved outcomes in the H/L group (p = 0.1141) (Figure 3). Although the finding was not conclusive, it highlights AI-HOPE-TP53’s functionality in executing multi-layered stratifications—simultaneously integrating tumor type, mutation status, and demographic metadata.

**Figure 3.**
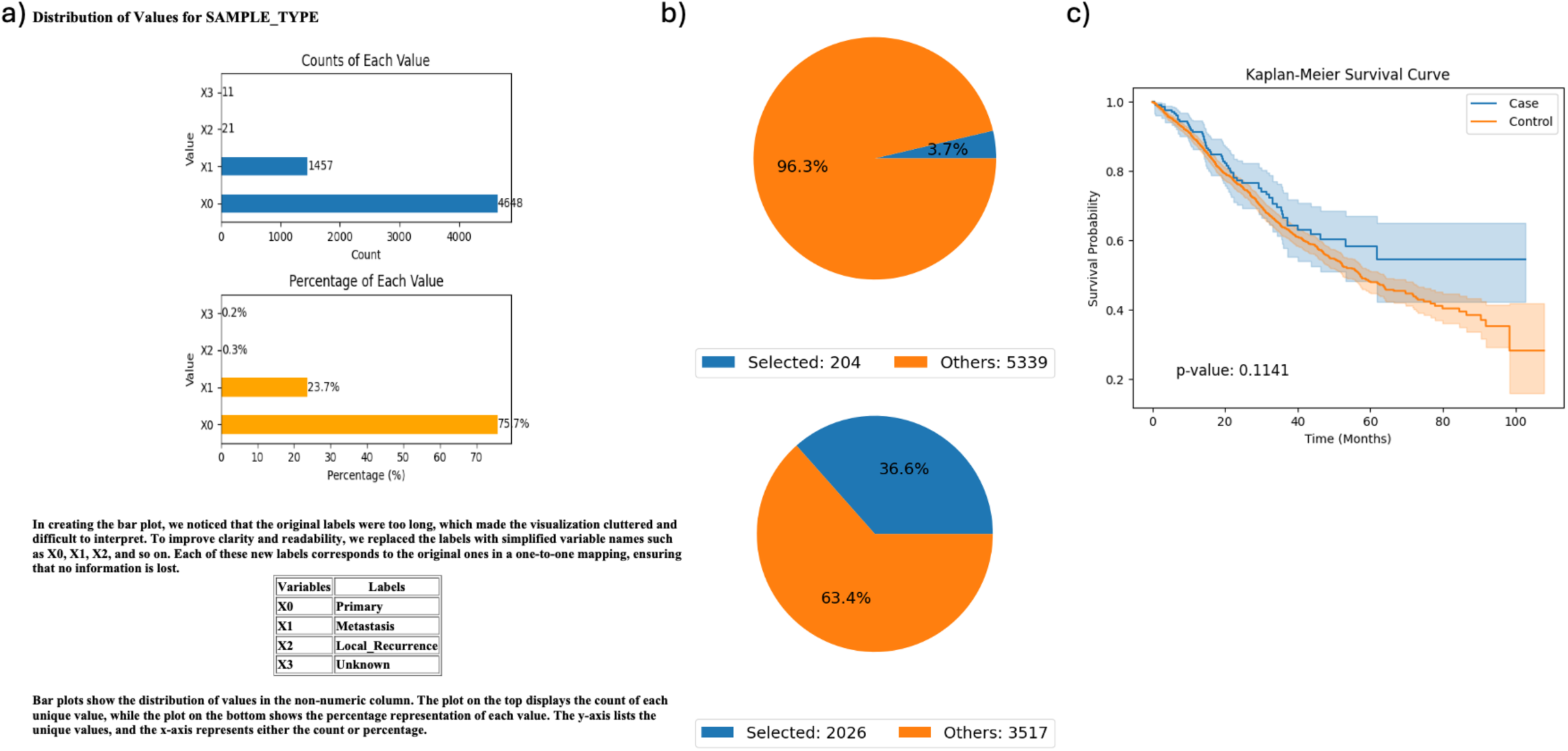
AI-HOPE-TP53 analysis of TP53-mutant primary colorectal cancer samples by ethnicity (Hispanic/Latino-H/L vs. Non-Hispanic White-NHW). This figure demonstrates the use of AI-HOPE-TP53 to assess survival outcomes in patients with TP53-mutated colorectal cancer (CRC), stratified by ethnicity and limited to primary tumor samples. a) Bar plots summarize the distribution of sample types across the dataset. The majority of samples are classified as primary tumors (X0), accounting for 75.7% of cases (n = 4,648), followed by metastatic samples (X1, 23.7%, n = 1,457). Labels were condensed for clarity, with X0 representing primary, X1 representing metastasis, and less common categories (e.g., local recurrence, unknown) grouped as X2 and X3. b) Pie charts visualize the cohort selection process after applying natural language query filters. The case cohort (H/L patients with TP53-mutated primary tumors) includes 204 samples (3.7% of the dataset), while the control cohort (NHW patients with TP53-mutated primary tumors) includes 2,026 samples (36.6%). These charts provide context for subgroup size and selection criteria. c) Kaplan-Meier survival curves compare overall survival between the two groups. While H/L patients with TP53-mutated primary CRC show a trend toward improved survival, the difference was not statistically significant (p = 0.1141). Shaded regions represent 95% confidence intervals.

### Exploratory analysis: Tumor subsite differences in ATM-mutant CRC

Survival differences by tumor subsite were investigated in CRC patients harboring ATM mutations. Patients with colon adenocarcinoma (n = 307) showed a trend toward poorer survival compared to those with rectal adenocarcinoma (n = 91), though this did not reach statistical significance (p = 0.3134) (Figure 4). This analysis highlights the platform’s ability to refine cohort definitions and enable subsite-specific comparisons in the context of pathway alterations.

**Figure 4.**
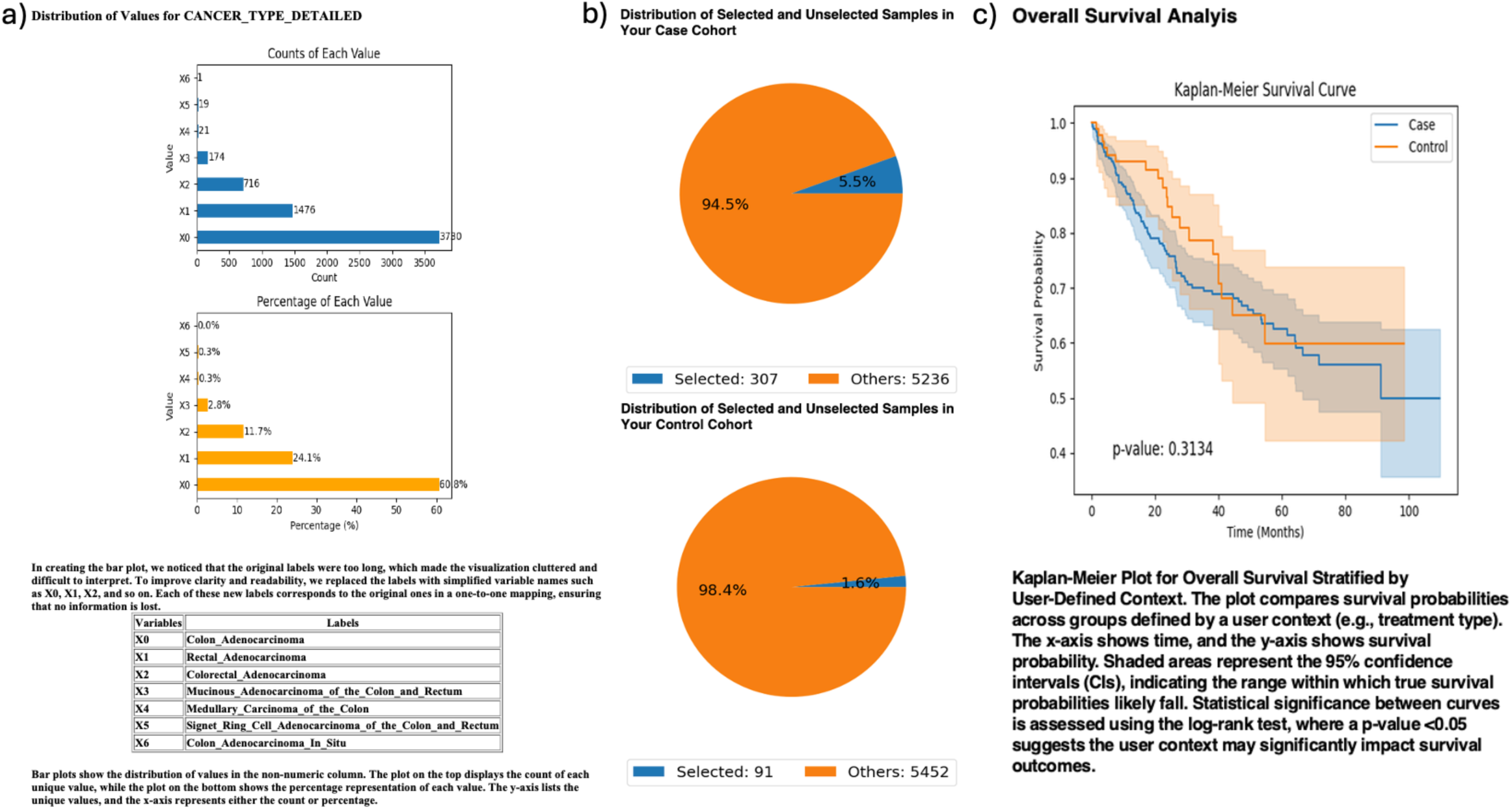
AI-HOPE-TP53 Analysis of ATM-Mutated Colorectal Cancer (CRC) Stratified by Tumor Subsite: Colon vs. Rectal Adenocarcinoma. This figure demonstrates the application of AI-HOPE-TP53 to evaluate survival outcomes among CRC patients harboring ATM mutations, stratified by tumor origin—colon adenocarcinoma (case) versus rectal adenocarcinoma (control). a) Bar plots display the distribution of detailed cancer subtypes across the dataset. Colon adenocarcinoma (X0) is the most prevalent subtype, comprising 60.6% of samples (n = 3,370), followed by rectal adenocarcinoma (X1, 24.1%, n = 1,341). Labels were simplified to improve clarity (e.g., X0 = Colon_Adenocarcinoma, X1 = Rectal_Adenocarcinoma). The top panel shows sample counts, while the lower panel shows their proportional representation. b) Pie charts highlight the selection of ATM-mutated samples for each group. The case cohort (colon adenocarcinoma with ATM mutations) includes 307 samples (5.5% of the dataset), and the control cohort (rectal adenocarcinoma with ATM mutations) includes 91 samples (1.6%). These distributions provide visual context for the relative representation of ATM-mutated cases across tumor subsites. c) Kaplan-Meier survival curves compare overall survival between ATM-mutated colon and rectal cancer cohorts. While the case group (colon adenocarcinoma) showed a trend toward poorer survival relative to the control group, the difference did not reach statistical significance (p = 0.3134). Confidence intervals are shown as shaded bands.

### Exploratory analysis: TP53-Mutated CRC and age-specific chemotherapy outcomes

AI-HOPE-TP53 was used to explore age-related survival differences among TP53- mutated CRC patients treated with FOLFOX chemotherapy (fluorouracil, leucovorin, oxaliplatin). Early-onset patients (<50 years; n = 1,000) demonstrated significantly improved survival compared to their late-onset counterparts (>50 years; n = 2,090), with a log-rank p-value of 0.0149 (Figure 5). Additionally, the early-onset group showed significantly greater representation of H/L patients (10.7%) compared to the late-onset group (5.65%). The corresponding odds ratio was 2.002 (95% CI: 1.524–2.632; p < 0.0001), indicating significant enrichment. This analysis underscores the platform’s ability to support intersectional modeling of molecular, demographic, and treatment factors.

**Figure 5.**
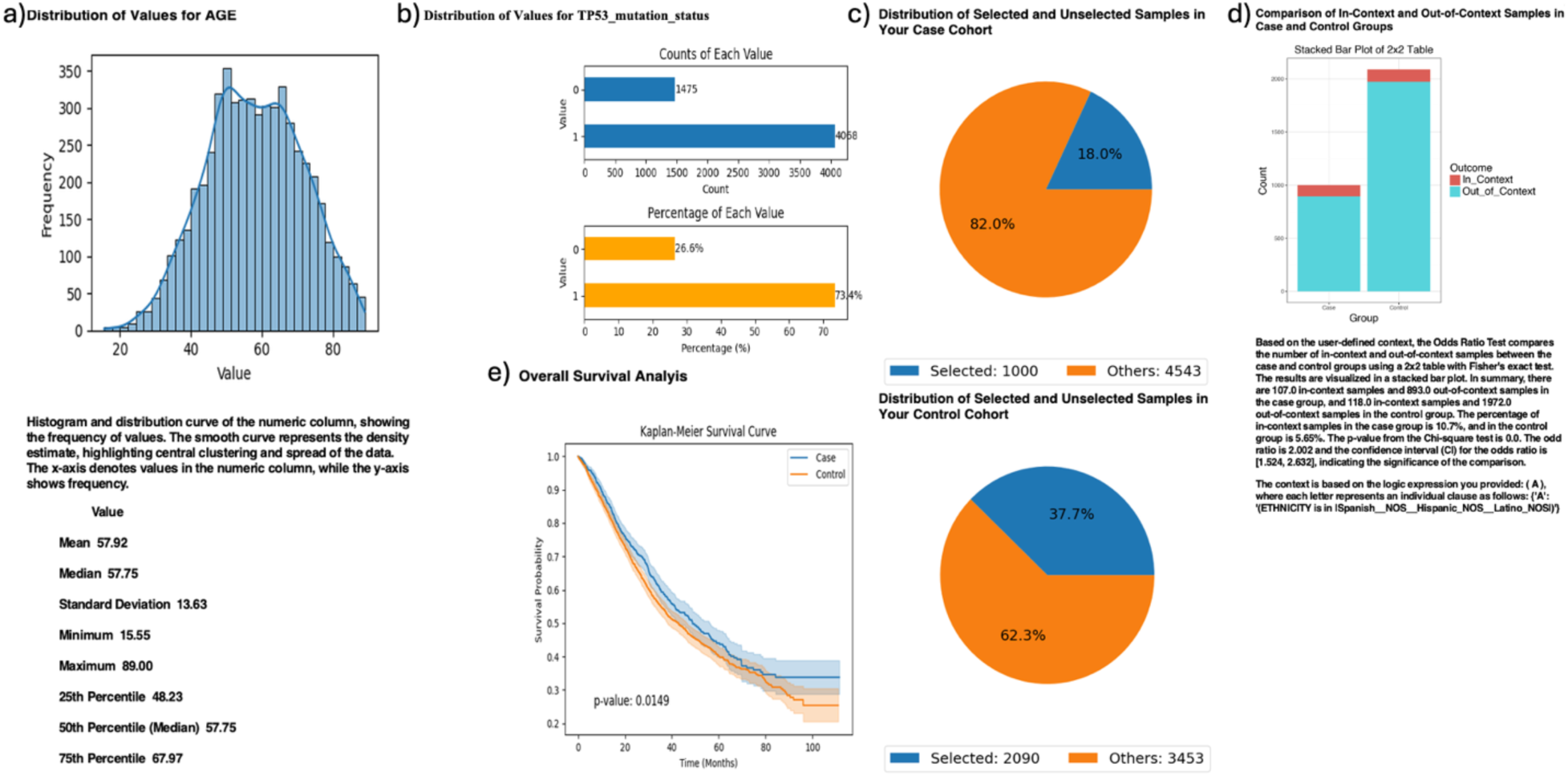
AI-HOPE-TP53 Analysis of TP53-Mutated Colorectal Cancer (CRC) Patients Treated with FOLFOX Chemotherapy, Stratified by Age and Evaluated for Hispanic/Latino (H/L) Representation. This figure demonstrates the use of AI-HOPE-TP53 to assess survival outcomes and ethnic enrichment among TP53-mutated CRC patients treated with the FOLFOX chemotherapy regimen (Fluorouracil, Leucovorin, and Oxaliplatin), comparing early-onset (<50 years) and late-onset (>50 years) cases. a) A histogram depicts the age distribution across the full dataset, with a mean of 57.92 years and a median of 57.75. This distribution contextualizes the cutoff for classifying early-onset CRC cases. b) Bar plots summarize the dataset-wide distribution of TP53 mutation status. TP53-mutated samples (X0) represent 26.5% of the cohort, while wild-type samples (X1) make up the remaining 73.5%. The top panel shows absolute counts, while the bottom displays proportional values. c) Pie charts illustrate the cohort selection process. The case cohort includes 1,000 early-onset CRC patients (age <50) with TP53 mutations treated with FOLFOX (18.0% of the dataset), while the control group consists of 2,090 late-onset CRC patients (age >50) with the same molecular and treatment profile (37.7%). d) A 2×2 odds ratio analysis evaluated the enrichment of H/L patients within the two cohorts. Among early-onset TP53-mutated patients treated with FOLFOX, 10.7% were H/L compared to 5.65% in the late-onset group. The calculated odds ratio was 2.002 (95% CI: 1.524–2.632; p < 0.0001), indicating statistically significant enrichment of H/L individuals in the early-onset group. e) Kaplan-Meier survival analysis compares overall survival between the case and control groups. Early-onset patients demonstrated significantly improved survival outcomes (p = 0.0149), with clear separation between curves and non-overlapping confidence intervals.

### Exploratory analysis: CHEK1 mutations and CRC stage

We next evaluated survival outcomes in CRC patients with CHEK1 mutations, comparing early-stage (Stage I–III) to advanced-stage (Stage IV) tumors. Although the early-stage group (n = 72) demonstrated a trend toward longer survival compared to the late-stage group (n = 6), the result was not statistically significant (p = 0.1592) (Figure S2). Despite small sample size, the analysis exemplifies how AI-HOPE-TP53 can be applied to rare mutation and stage-specific subgroups—domains often underpowered in traditional analysis pipelines.

### Exploratory analysis: Gender-based outcomes in TP53 pathway–altered CRC patients receiving FOLFOX

Finally, we evaluated gender differences in survival among CRC patients with TP53 pathway alterations receiving FOLFOX chemotherapy. Although females (n = 2,122) showed a trend toward improved survival relative to males (n = 2,470), the result was not statistically significant (p = 0.0998). Notably, the odds ratio analysis revealed that females were underrepresented among FOLFOX-treated TP53-altered cases (OR = 0.845, 95% CI: 0.741–0.963; p = 0.0138) (Figure S3), raising important questions regarding treatment access or selection bias across genders. AI-HOPE-TP53 was able to uncover these discrepancies through contextual query handling and pathway-focused filtering. Together, these validation and exploratory analyses underscore the flexibility, robustness, and translational relevance of AI-HOPE-TP53. The system successfully recapitulated previously reported associations while enabling new hypothesis generation in underexplored domains, including ethnicity-associated survival disproportionate health burdens, age-stratified therapy outcomes, and rare mutation-specific subgroup dynamics. By integrating conversational artificial intelligence with clinical-genomic bioinformatics, AI-HOPE-TP53 empowers researchers and clinicians to interrogate the TP53 signaling axis in colorectal cancer across demographic, molecular, and treatment dimensions—enhancing precision oncology efforts with equity-centered insights.

## Discussion

The development and application of AI-HOPE-TP53 represent a significant advancement in the field of precision oncology, particularly for the pathway-specific and disparity-aware investigation of colorectal cancer (CRC). By combining the capabilities of large language models with harmonized clinical-genomic datasets, AI-HOPE-TP53 addresses key limitations in current analytic tools—most notably, the inability to dynamically stratify patient cohorts across intersecting biological and sociodemographic variables using natural language input.

Our validation analyses confirm the platform’s ability to replicate established clinical-genomic associations. In particular, AI-HOPE-TP53 detected a consistent trend toward higher TP53 pathway alteration frequencies in early-onset colorectal cancer (EOCRC) among H/L compared to NHW patients. Although these trends did not reach statistical significance (Figures 2 and S1), the observed effect sizes were directionally aligned with prior reports on the enrichment of genomic instability and aggressive molecular features in EOCRC within underrepresented groups. These findings reinforce the hypothesis that TP53 pathway dysregulation may contribute to the disparate burden of CRC in H/L populations and support further investigation in larger and more diverse cohorts.

Exploratory analyses revealed the broader utility of AI-HOPE-TP53 in generating novel insights. For example, Kaplan-Meier survival modeling indicated a potential ethnicity-specific survival advantage among H/L patients with TP53-mutated primary tumors (Figure 3). Additionally, our results demonstrated the platform’s capacity to perform refined subsite-specific survival comparisons in ATM-mutated CRC (Figure 4), and to evaluate age-dependent outcomes among FOLFOX-treated, TP53-mutated patients. Notably, early-onset patients showed both improved survival and a significantly higher proportion of H/L representation (Figure 5), highlighting a possible interaction between molecular alterations, treatment response, and population with disproportionate health burdens that warrants further mechanistic study.

AI-HOPE-TP53 also enabled the dissection of less commonly studied subgroups, such as CRC patients with CHEK1 mutations stratified by tumor stage (Figure S2). Despite limited sample sizes, the system provided interpretable survival analyses and contextual odds ratio assessments, underscoring its value for hypothesis generation in rare mutation contexts. Furthermore, the detection of gender-based underrepresentation in FOLFOX-treated TP53-altered cases (Figure S3) suggests potential differences in treatment access or eligibility that are not always captured in standard data workflows.

These findings emphasize the importance of equity-informed analytic capabilities in precision oncology platforms. Taken together, these results demonstrate that AI-HOPE-TP53 is more than a data analysis tool—it is a hypothesis-generation engine capable of democratizing access to complex clinical-genomic analyses. Its natural language–driven interface lowers the barrier for non-computational researchers and clinicians, enabling precision oncology studies that are both scalable and inclusive. Importantly, the platform’s modular architecture and retrieval-augmented generation (RAG) system provide robust safeguards against misinterpretation and ensure reproducibility across use cases.

Nonetheless, several limitations should be acknowledged. First, although AI-HOPE-TP53 is validated on publicly available datasets such as cBioPortal, communities with disproportionate health burden in cohort representation may limit generalizability to broader populations. Second, while trends identified in this study suggest biologically meaningful associations, many did not reach statistical significance—highlighting the need for further validation in larger, prospectively collected datasets. Finally, future iterations of the platform should integrate additional omic layers (e.g., transcriptomics, proteomics) and support real-world clinical data inputs to improve generalizability and clinical utility.

## Conclusion

In conclusion, AI-HOPE-TP53 establishes a new paradigm for pathway-centric, population-aware precision oncology. By enabling natural language–based interrogation of TP53-related molecular events in CRC, the system empowers translational researchers to uncover both established and novel insights—accelerating the path from data to discovery in cancer research.

## Data Availability

All data used in the present study is publicly available at https://www.cbioportal.org/ and https://genie.cbioportal.org. Additional data can be provided upon reasonable request to the authors.

---

**Figure S1.**
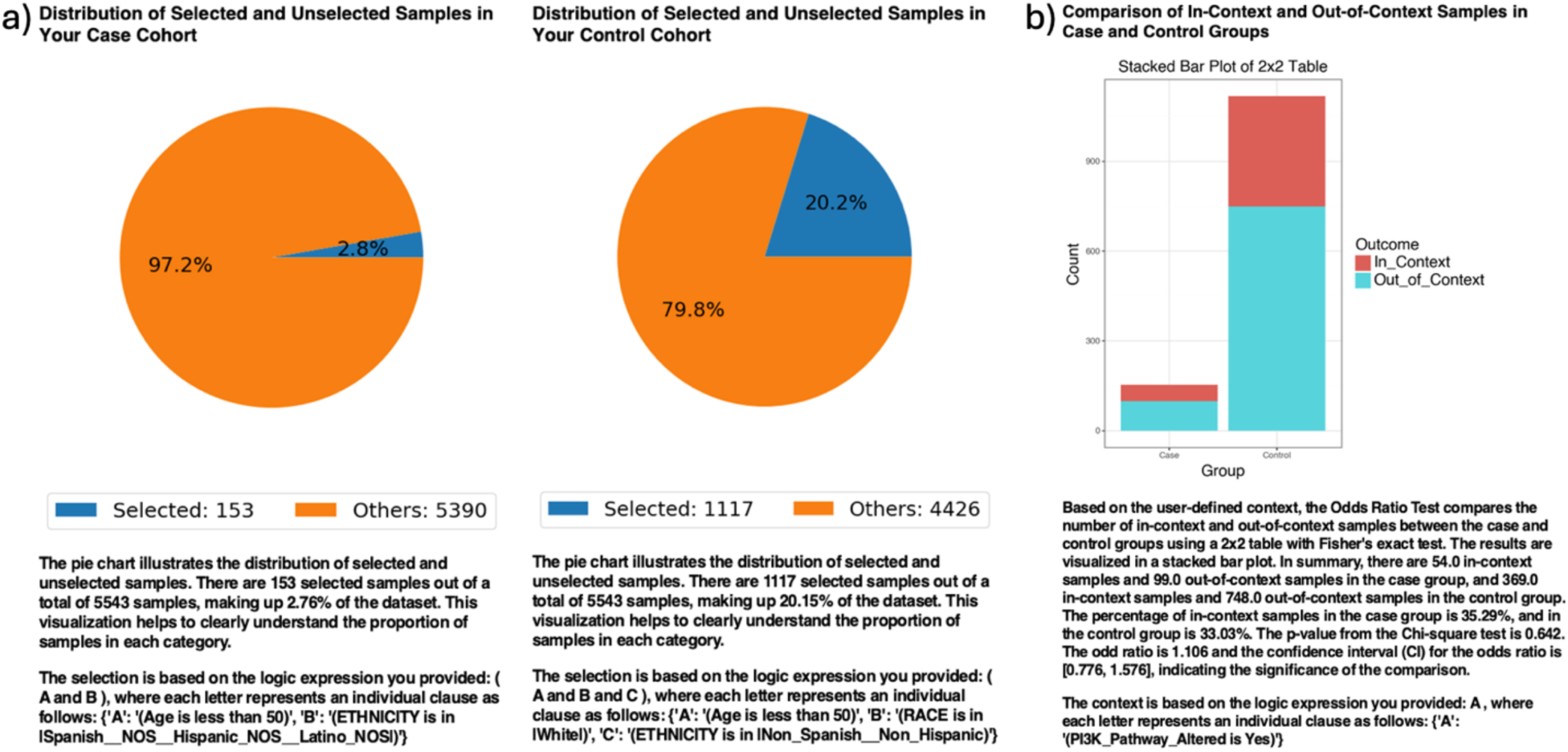
AI-HOPE-TP53 analysis of TP53 pathway alterations in early-onset colorectal cancer (EOCRC) among Hispanic/Latino (H/L) vs. Non-Hispanic White (NHW) patients. a) Pie charts illustrate the proportion of selected samples after applying natural language query filters. The case cohort (H/L) includes 153 EOCRC patients under age 50 with H/L ethnicity, representing 2.8% of the total dataset. The control cohort (NHW) includes 1,117 EOCRC patients under age 50 who are NHW, accounting for 20.2% of the queried population. b) A 2×2 odds ratio analysis compares the frequency of TP53 pathway alterations between the two cohorts. The stacked bar plot depicts the distribution of in-context (alteration present) and out-of-context (alteration absent) samples for both groups. TP53 pathway alterations were observed in 90.2% of H/L samples and 85.05% of NHW samples. The resulting odds ratio was 1.62 (95% CI: 0.926–2.825), with a p-value of 0.114, indicating a non-significant trend toward higher TP53 alteration frequency in H/L patients.

**Figure S2.**
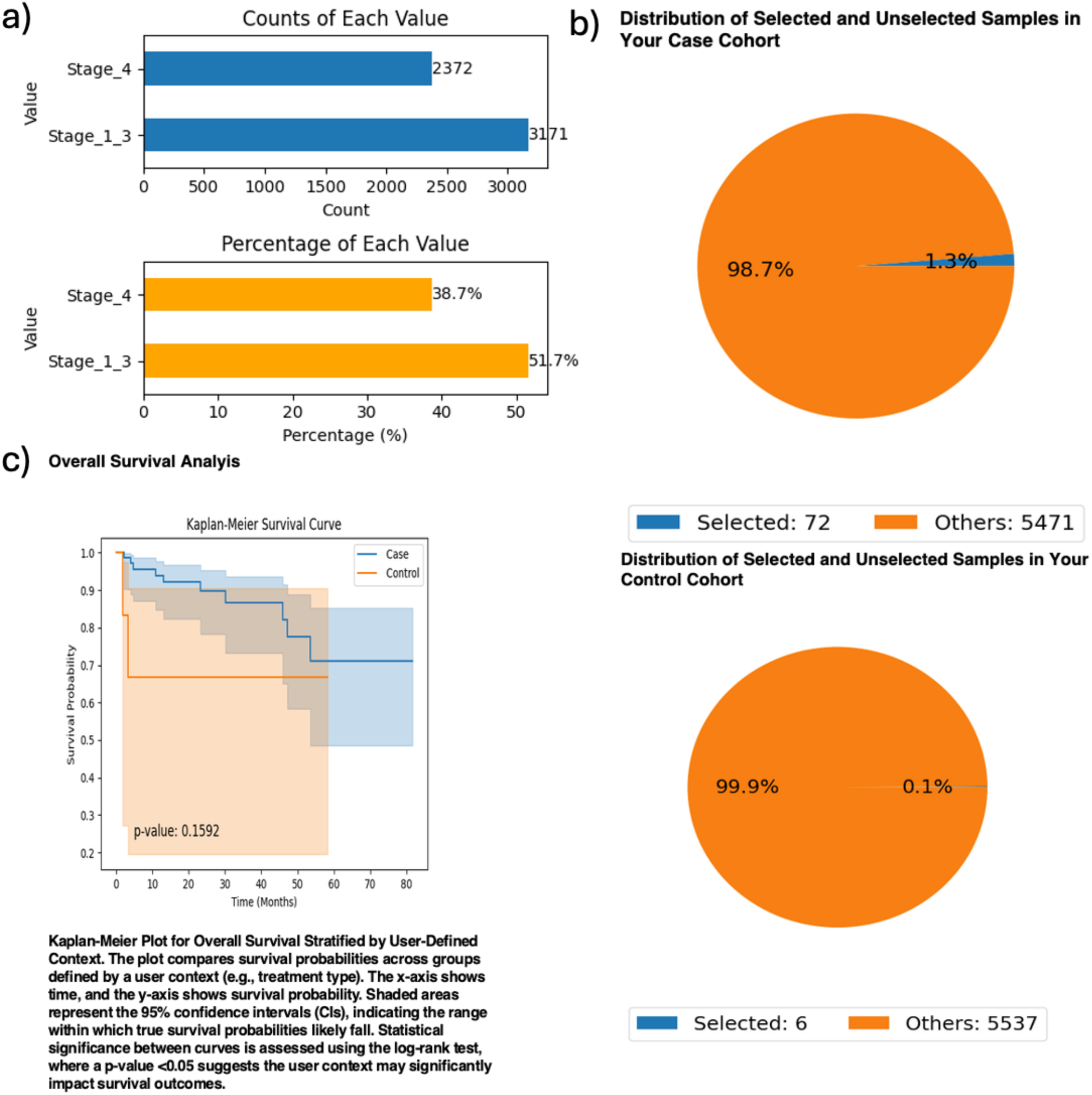
AI-HOPE-TP53 Analysis of CHEK1-Mutant Colorectal Cancer (CRC) Patients by Tumor Stage (Stage I–III vs. Stage IV). This figure illustrates the application of AI-HOPE-TP53 to evaluate survival outcomes among CRC patients harboring CHEK1 mutations, stratified by disease stage. The case cohort includes patients with early-stage tumors (Stage I–III), while the control cohort comprises patients with advanced-stage tumors (Stage IV). a) Bar plots display the dataset-wide distribution of clinical stage categories. Stage I–III cases (labeled as “Stage_1_3”) represent 51.7% of the total dataset (n = 3,171), while Stage IV cases account for 38.7% (n = 2,372). The top panel shows absolute sample counts, and the bottom panel shows proportional representation. b) Pie charts illustrate the cohort selection process based on natural language query filters. The early-stage CHEK1-mutated cohort includes 72 selected samples (1.3% of the dataset), while the advanced-stage CHEK1-mutated control group includes 6 samples (0.1%). These visuals highlight the rarity of CHEK1 mutations in stage-stratified CRC subgroups. c) Kaplan-Meier survival analysis compares overall survival between early-stage and late-stage CHEK1-mutated CRC patients. Although early-stage patients show a trend toward improved survival, the difference was not statistically significant (p = 0.1592). Shaded regions represent the 95% confidence intervals.

**Figure S3.**
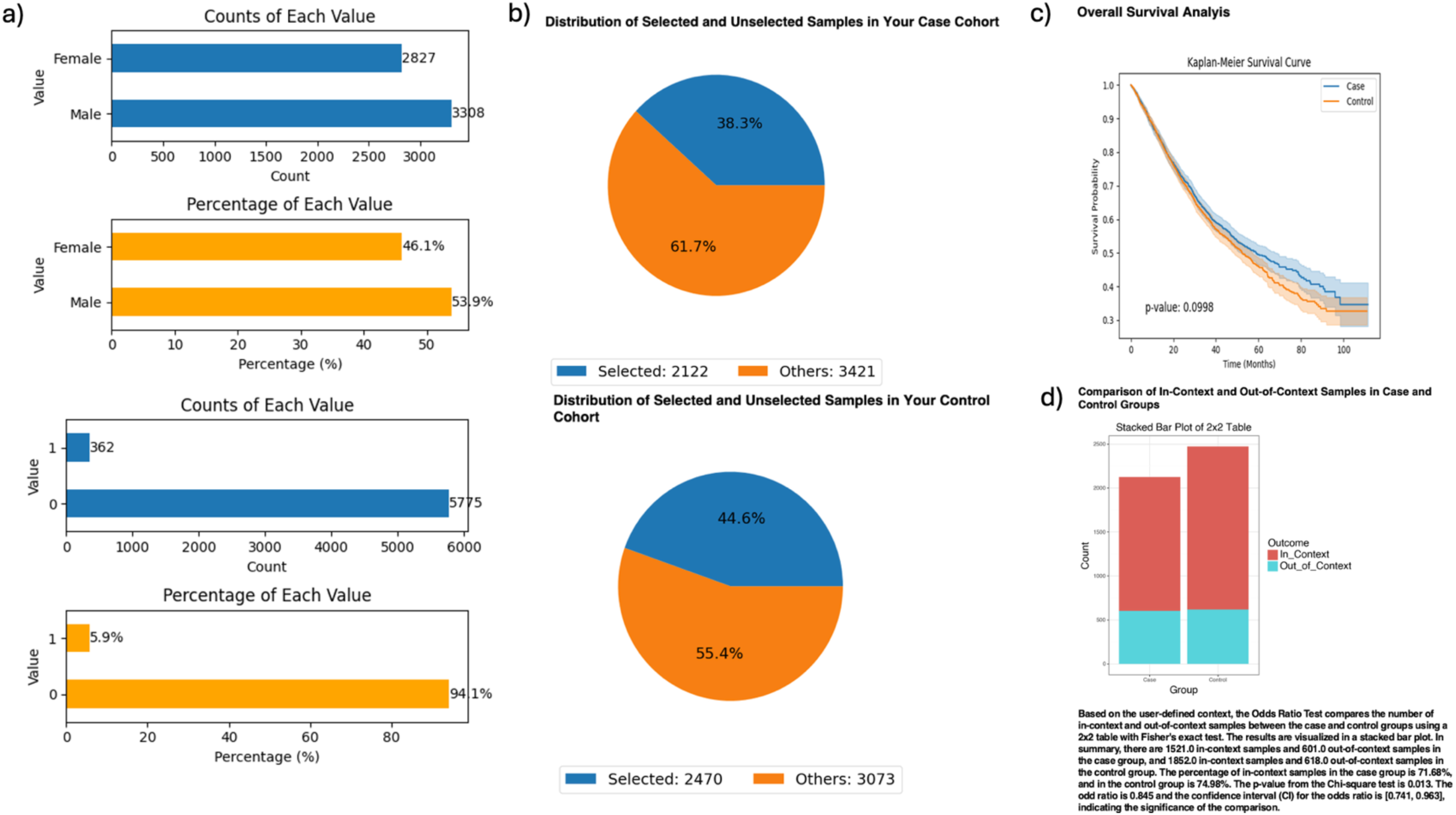
AI-HOPE-TP53 Analysis of Gender-Based Survival in TP53 Pathway– Altered Colorectal Cancer (CRC) Patients Receiving FOLFOX Chemotherapy. This figure demonstrates the use of AI-HOPE-TP53 to analyze sex-based differences in clinical outcomes among CRC patients with TP53 pathway alterations, contextualized by receipt of standard FOLFOX chemotherapy (Fluorouracil, Leucovorin, Oxaliplatin). a) Bar plots summarize baseline cohort characteristics. The top panel illustrates gender distribution across the dataset, with males (n = 3,398; 53.9%) slightly outnumbering females (n = 2,827; 46.1%). The bottom charts display counts and proportions of TP53 pathway–altered samples receiving FOLFOX treatment (Yes = 362, No = 5,775), highlighting the relative rarity of this mutation-treatment context. b) Pie charts depict the number of selected samples after applying the user-defined natural language query. The case cohort (females with TP53 pathway alterations) includes 2,122 samples (38.3%), while the control cohort (males with TP53 pathway alterations) includes 2,470 samples (44.6%). These visuals reflect near-equivalent representation and support gender-stratified comparative analysis. c) Kaplan-Meier survival analysis compares overall survival between female and male patients with TP53 pathway–altered CRC undergoing FOLFOX chemotherapy. Although the curves suggest a trend toward improved survival in female patients, the difference did not reach statistical significance (p = 0.0998). Confidence intervals overlap throughout the follow-up period. d) A 2×2 odds ratio analysis contextualized by FOLFOX exposure evaluates the enrichment of in-context samples (defined by gender and TP53 pathway mutation) between cohorts. The stacked bar plot shows that 71.68% of female samples and 74.96% of male samples met the full query context. The odds ratio was 0.845 (95% CI: [0.741, 0.963]; p = 0.0138), suggesting a statistically significant underrepresentation of FOLFOX-treated TP53 pathway–altered female patients relative to their male counterparts.

## Notes

### Competing Interest Statement

The authors have declared no competing interest.

### Funding Statement

This study was funded by the National Cancer Institute NCI award number U2CCA252971

### Author Declarations

The source data used in this study were publicly available before the initiation of the study and can be accessed through cBioPortal for Cancer Genomics at https://www.cbioportal.org/ and the GENIE Project (AACR Project GENIE cBioPortal) at https://genie.cbioportal.org. Additional data may be provided upon reasonable request to the authors.

### Summary of Updates

This version of the manuscript has been revised to update the title; the content of the manuscript remains unchanged.

